# Colorectal cancer is associated with the presence of cancer driver mutations in normal colon

**DOI:** 10.1101/2021.10.11.21264780

**Authors:** Julia Matas, Brendan Kohrn, Jeanne Fredrickson, Kelly Carter, Ming Yu, Ting Wang, Xianyong Gui, Thierry Soussi, Victor Moreno, William M. Grady, Miguel A. Peinado, Rosa Ana Risques

## Abstract

While somatic mutations in colorectal cancer (CRC) are well characterized, little is known about the accumulation of cancer mutations in the normal colon prior to cancer. Here we have developed and applied an ultra-sensitive, single-molecule mutational test based on CRISPR-DS technology, which enables mutation detection at extremely low frequency (<0.001) in normal colon from patients with and without CRC. We found oncogenic *KRAS* mutations in the normal colon of about one third of patients with CRC but in none of the patients without CRC. Patients with CRC also carried more *TP53* mutations than patients without cancer, and these mutations were more pathogenic and formed larger clones, especially in patients with early onset CRC. Most mutations in normal colon were different from the driver mutations in tumors suggesting that the occurrence of independent clones with pathogenic *KRAS* and *TP53* mutations is a common event in the colon of individuals that develop CRC.

**SIGNIFICANCE:** Our results suggest a prevalent process of somatic mutation and evolution in the normal colon of patients with CRC, which can be detected by ultra-sensitive sequencing of driver genes and potentially employed clinically for CRC risk prediction.

## INTRODUCTION

Colorectal cancers, like many other solid tumors, form through a process of somatic evolution that takes decades (1). This fact and our current understanding of the molecular pathogenesis of colorectal adenomas and adenocarcinoma imply that clones primed with cancer mutations exist within histologically normal tissue for years prior to cancer diagnosis. Providing convincing evidence of this proposed phenomenon is of critical importance for early cancer detection, risk prediction, and prevention (2,3). However, the robust identification of these mutant clones has been challenging due to the lack of methods able to reliably detect small subsets of mutant cells within morphologically normal tissue.

Two approaches to identify somatic mutations in normal tissue consist of either reducing the sample size to very small or microscopic regions in which mutant clones represent a sizable proportion of cells (4), or of expanding single-cells in vitro to larger clones amenable for next-generation sequencing (NGS) (5). These approaches have been applied to the characterization of somatic mutations in normal tissues including skin (6,7), esophagus (8,9), endometrium (10,11), bladder (12,13), colon (14,15), and others (16,17). Many of these studies have revealed an unexpected abundance of cancer driver mutations in histologically normal tissue (18,19) and are rapidly expanding our understanding of the role of somatic evolution in human aging (20,21). However, little is known about how this process differs in individuals with and without cancer, and whether this knowledge can be harnessed to improve cancer prediction and prevention. A main challenge relies on the fact that these approaches require the analysis of a large number of samples per individual and are very labor-intensive, precluding large cohort studies and translational applications.

An alternative approach to detect low frequency somatic mutations within normal tissue consists of performing ultra-deep sequencing using high-accuracy NGS methods such as duplex sequencing (DS) (22). DS employs double-stranded molecular tags, which enable error correction by consensus sequence independently in each DNA strand, effectively decreasing the error rate of sequencing from 10^−3^ to <10^−7^ (23). Because each duplex read corresponds to an original DNA molecule, this method enables the detection of single mutant DNA molecules among thousands of non-mutant genomes, thus providing extreme resolution to identify mutant cells in normal tissue by analyzing a single biopsy. The trade-off for this high-throughput, high-sensitive approach for mutation detection is that it requires large sequencing capabilities and therefore is mostly suitable for small target regions. We have used DS to perform ultra-deep sequencing of *TP53* in normal gynecological tissues across the human lifespan, revealing a progressive enrichment of *TP53* pathogenic mutations with older age (24). We have also demonstrated the presence of cancer driver *TP53* mutations in the peritoneal fluid (25), uterine lavage (24) and Pap test DNA (26) of women with and without ovarian cancer. Women with cancer tended to have higher *TP53* mutation burden (25,26), suggesting increased *TP53* somatic evolution in association with cancer progression.

Colorectal cancer is the second most common cause of cancer death in the USA and its incidence has been increasing in individuals under 50 years of age (27). The need for better biomarkers for CRC prediction, coupled with easy access to normal tissue via colonoscopy, makes this cancer type especially suitable to study the potential of clonal expansion detection in histologically normal tissue to identify early cancer progression. Our goal was to investigate whether mutations in common CRC genes (*TP53, KRAS, PIK3CA* and *BRAF*) could be detected by ultra-deep sequencing (>1,000x) in single, histologically normal colon biopsies and to determine whether they were more frequent in individuals with CRC than in those who are cancer-free. These genes were selected because, together with *APC*, they constitute the 5 most frequently mutated genes in CRC. However, in contrast to *APC*, they accumulate mutations in localized hotspot regions, thus providing excellent targets for the development of ultra-sensitive sequencing tests for early cancer detection. We used a version of DS called CRISPR-DS (28), which employs CRISPR-based target enrichment to increase library preparation efficiency for small target panels. We demonstrate the feasibility of our approach to detect low frequency cancer driver mutations in normal colon, and the potential clinical value of these mutations as CRC risk markers. Our results suggest that this approach could be useful for CRC risk prediction in younger patients, who constitute a rapidly increasing subset of the population at risk (29).

## RESULTS

### CRISPR-DS enables ultra-sensitive detection of mutations in normal colon and CRC cell lines

We used CRISPR-DS to perform ultra-deep sequencing (minimum 1,000x, mean ∼2,500x) of DNA from the normal colonic epithelium of 47 individuals: 24 without CRC and 23 with CRC (Fig. 1A and Methods). Half of the patients with CRC were 50 years of age or younger to allow investigation of the role of somatic mutation load in individuals with early age of onset of CRC. The clinico-pathological characteristics of the individuals are detailed in Supplementary Table S1. We analyzed normal left colon epithelium in individuals without CRC and normal epithelium distant from tumor (>10cm in most cases) in individuals with CRC.

**Figure 1.**
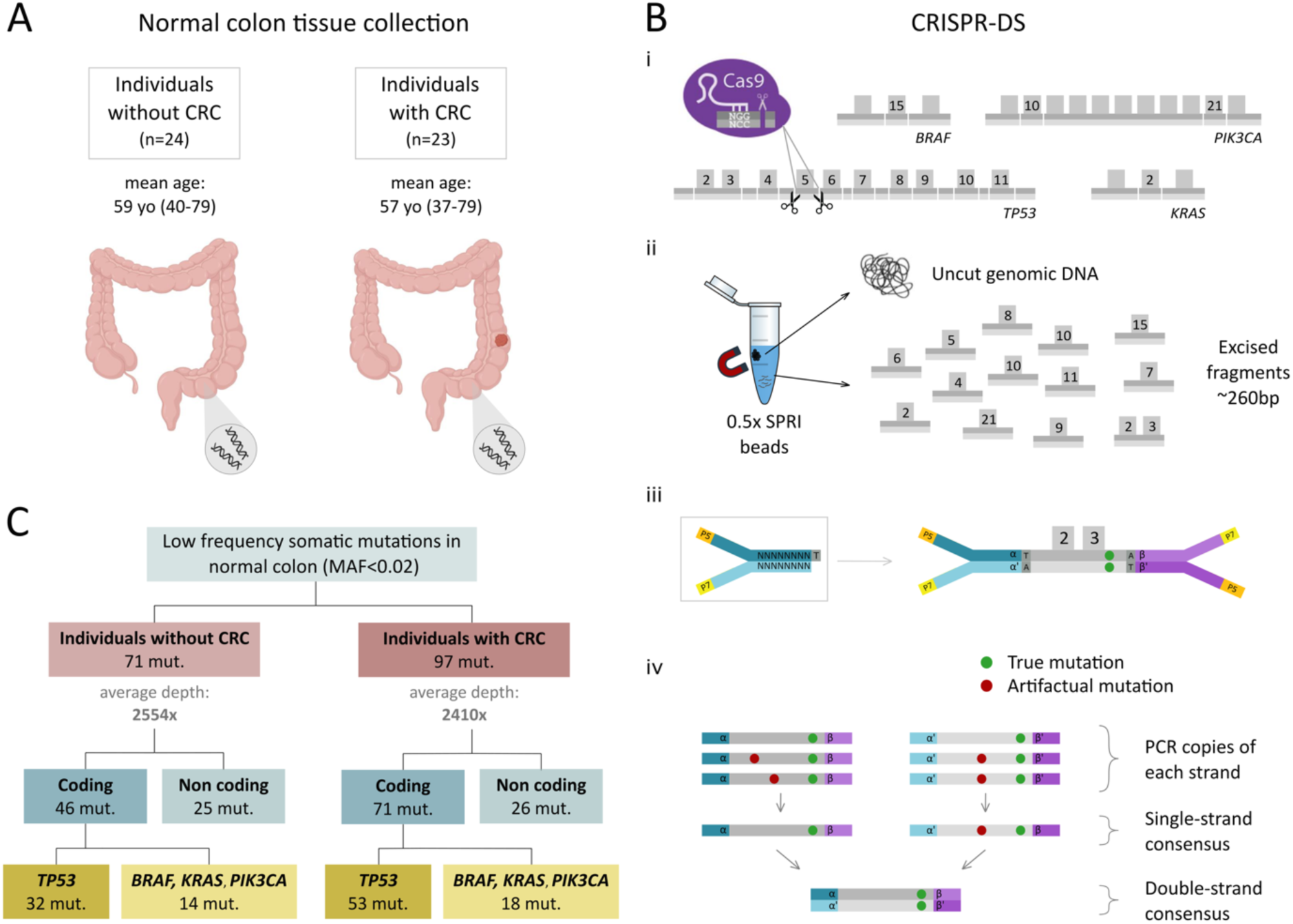
CRISPR-DS enables ultra-sensitive detection of cancer gene mutations in normal colon samples. **A**. Normal colon biopsies were procured from individuals with and without CRC. **B**. CRISPR-DS. (i) CRISPR-Cas9 guides were designed to target common oncogenic mutation regions in *BRAF, PIK3CA* and *KRAS* as well as the full coding region of *TP53* in fragments of ∼260bp. (ii) Size selection with SPRI beads was used to enrich for excised fragments. (iii) Excised fragments were ligated to adapters with random double-stranded molecular tags. (iv) The generation of single-strand and double-strand consensus sequences from reads sharing the same molecular tags enables the elimination of artifactual mutations. Adapted from Nachmanson *et al* (ref. 28). **C**. Low frequency somatic mutations in cancer genes are identified in normal colon from patients with and without CRC.

CRISPR-DS enables ultra-accurate deep-sequencing of selected genomic regions, which are excised with CRISPR-Cas9 in fragments of predetermined size. These fragments are then size-selected for target enrichment prior to library preparation (28). This process eliminates problems associated with DNA sonication (30) and increases sequencing efficiency (28). We adapted this method to digest in ∼260bp fragments the exons that carry common oncogenic mutations in *BRAF, KRAS* and *PIK3CA* and the full coding region of *TP53* (Fig. 1B and Methods). The CRISPR RNA (crRNA) sequences for CRISPR-Cas9 digestion and the probes for hybridization capture are indicated in Supplementary Tables S2 and S3, respectively. Duplex adapters containing 8bp random molecular tags were used to uniquely label each DNA molecule to enable double-strand error correction as previously described (22,28) (Fig. 1B, Methods).

We first demonstrated the reproducibility and sensitivity of the assay by deep sequencing DNA from 3 common CRC cell lines with driver mutations in our target genes (HCT116, SW480 and HT29) (Supplementary Methods). All the expected driver mutations were identified in addition to multiple low frequency (<0.1) mutations in HCT116 and several low frequency mutations in SW480, in agreement with the known levels of single nucleotide variants in these cell lines (31) (Supplementary Fig. S1A). An independent technical replicate experiment identified not only the expected driver mutations but also all the expected mutations with Mutant Allele Frequency (MAF) as low as 0.001 and a subset of the very rare mutations below 0.001, despite the decreased likelihood of resampling of a rare event (Supplementary Fig. S1A). These results demonstrate the high sensitivity and reproducibility of the assay even at 0.001 MAF. To further demonstrate sensitivity and accuracy in an independent experiment, we spiked DNA from HT29 into DNA from HCT116 at 3 different ratios (1:10, 1:20, 1:100). The two driver HT29 mutations were observed at the expected frequencies in the 3 mixes even when present at low level (0.01 and 0.003) (Supplementary Fig. S1B).

We then used CRISPR-DS to sequence the normal colon of individuals with and without CRC. While the mean duplex depth across samples was variable, all samples reached a minimum of 1,000x duplex depth and the average depth for both groups of patients was similar (Fig. 1C). Overall, CRISPR-DS yielded a total of 404M duplex nucleotides, with 227M in coding regions and 177M in non-coding regions. A total of 168 mutations were identified: 117 coding and 51 non-coding (Fig. 1C). All these mutations had low MAF (<0.02). As in prior studies (24-26), the number of mutations tended to increase with the number of duplex nucleotides sequenced (Supplementary Fig. S2). To correct for this effect, sample comparisons were made based on mutation frequencies, which were calculated as the number of mutations in a given region (e.g., coding exons) divided by the total number of duplex nucleotides sequenced in that region (Methods). Mutation counts and corresponding mutation frequencies for each sample are shown in Supplementary Table S4.

### Individuals with CRC carry a higher frequency of coding mutations than individuals without cancer in a non-age dependent manner

We first compared coding and non-coding mutation frequencies in the normal colon of patients with and without CRC. Patients with CRC had a significantly higher coding mutation frequency in normal colon than patients without cancer (t-test p=0.005, Fig. 2A) even when separating the patients by early and late stage CRC (Supplementary Fig. S3). Non-coding mutation frequency, however, was similar in patients with and without CRC (Fig. 2A). Interestingly, the non-coding mutation frequency significantly increased with age (Spearman’s correlation p=0.024), but this trend was not observed for the coding mutation frequency (Fig. 2B). In addition, higher non-coding mutation frequency correlated with advanced epigenetic age in the normal colon as measured by the Horvath clock, the PhenoAge clock, and the EpiTOC clock, which are well-established measurements of epigenetic aging (32,33) (Fig. 2C). Coding mutations did not associate with lower or higher epigenetic age determined by these clocks. These results indicate that while intronic, non-functional mutations accumulate with chronological and biological aging in the normal colon, coding mutations in target driver genes exceed the age-related background level, especially in patients that develop CRC.

**Figure 2.**
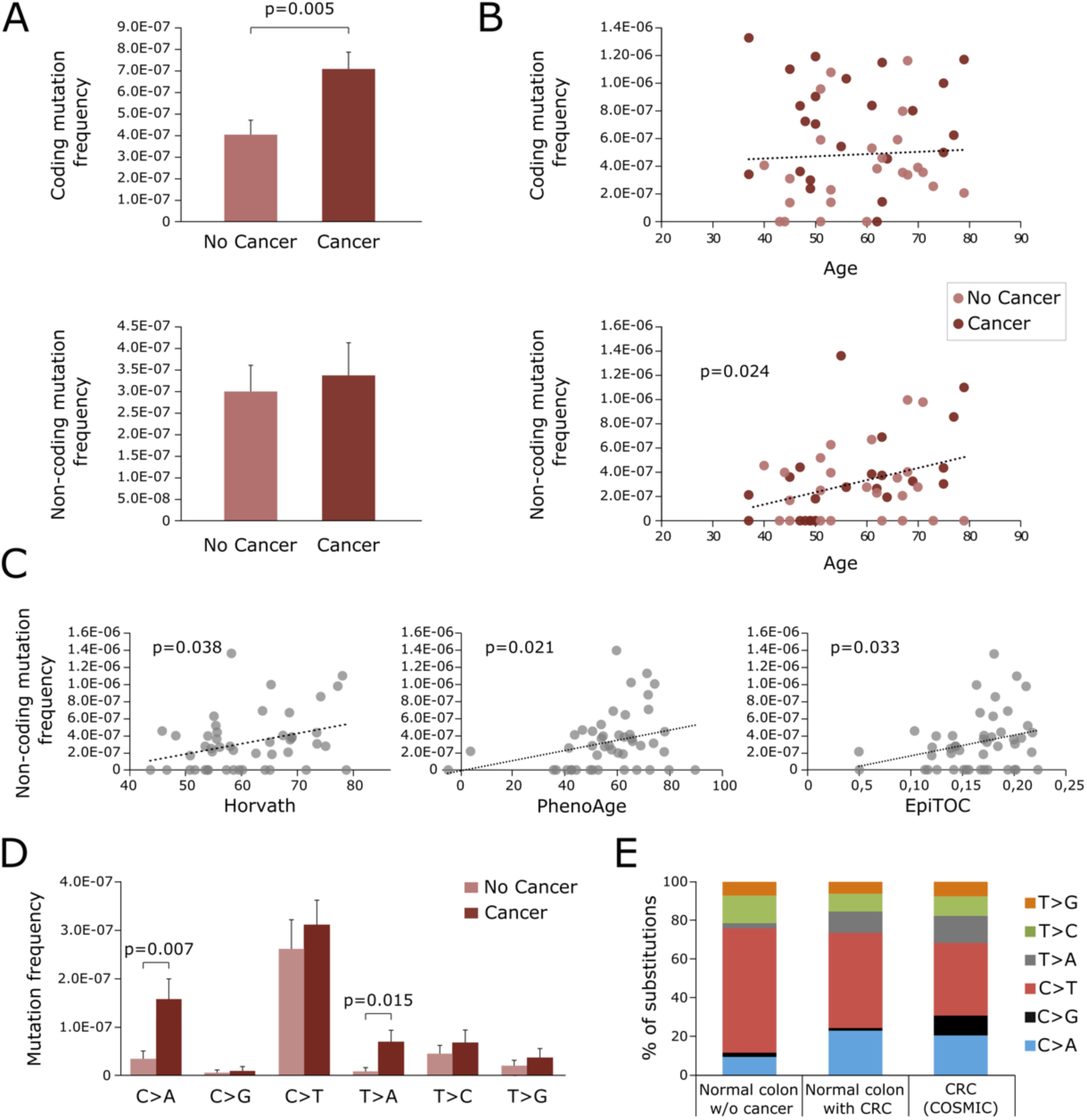
Normal colon of patients with CRC has higher, not age-related, coding mutation frequency and a mutation spectrum similar to cancers. **A**. Coding and non-coding mutation frequency in normal colon from individuals with and without cancer. Mutation frequency is calculated as the number of mutations divided by the total number of duplex nucleotides sequenced in the coding or non-coding target regions, respectively. P-value corresponds to t-test. Error bars represent standard error of the mean. **B**. Coding and non-coding mutation frequency and its correlation with age. P-value corresponds to Spearman’s correlation. **C**. Non-coding mutation frequency correlation with Horvath, PhenoAge and EpiTOC epigenetic clocks. P-values correspond to Spearman’s correlation. **D**. Frequency of coding mutation by substitution type compared between normal colon of individuals with and without cancer. P-values correspond to t-tests. Error bars represent standard error of the mean. **E**. Mutation spectrum compared between coding nucleotide substitutions from normal colon of individuals without CRC (n=42), with CRC (n=65), and the CRC COSMIC database (n=70,525). Only significant p-values are displayed.

To further explore the nature of the coding mutations present in the normal colon of patients with and without cancer, we classified them by mutational spectrum (C>A, C>G, C>T, T>A, T>C, T>G). While all types of mutations were more frequent in patients with cancer, two types were significantly overrepresented: C>A (t-test p=0.007) and T>A (t-test p=0.015) (Fig. 2D). These two types of mutations are also enriched in CRC based on the Catalogue of Somatic Mutations in Cancer (COSMIC) database (34) (Fig. 2E). These results indicate that the normal colon in individuals with CRC not only carries more coding mutations, but these mutations are more similar to those found in tumors.

### *KRAS* and *TP53* driver mutations are abundant in the colon of patients with CRC

We then explored the distribution of coding mutations by gene (Fig. 3A-C). For *BRAF, PIK3CA* and *KRAS*, we deep sequenced the exons with CRC mutation hotspots according to the COSMIC database. We found several mutations in *BRAF* and *PIK3CA*, but none of the *BRAF* mutations (0/7) corresponded to the canonical hotspot V600E mutation and only two of the *PIK3CA* mutations (2/12) corresponded to the 3 most common *PIK3CA* mutations in CRC, which account for >50% of CRC *PIK3CA* mutations according to COSMIC (34) (Fig. 3A-B, Methods, and Supplementary Table S5). In contrast, 7 out of 13 *KRAS* mutations identified (54%) corresponded to oncogenic hotspot mutations in codons 12 or 13 (Fig. 3C). Remarkably, these 7 oncogenic *KRAS* mutations were all identified in normal colon from individuals with CRC. Overall, 30% of patients with CRC carried a *KRAS* hotspot mutation in normal colon compared to none of the patients without CRC (Fig. 3D, Pearson Chi-Square p=0.003).

**Figure 3.**
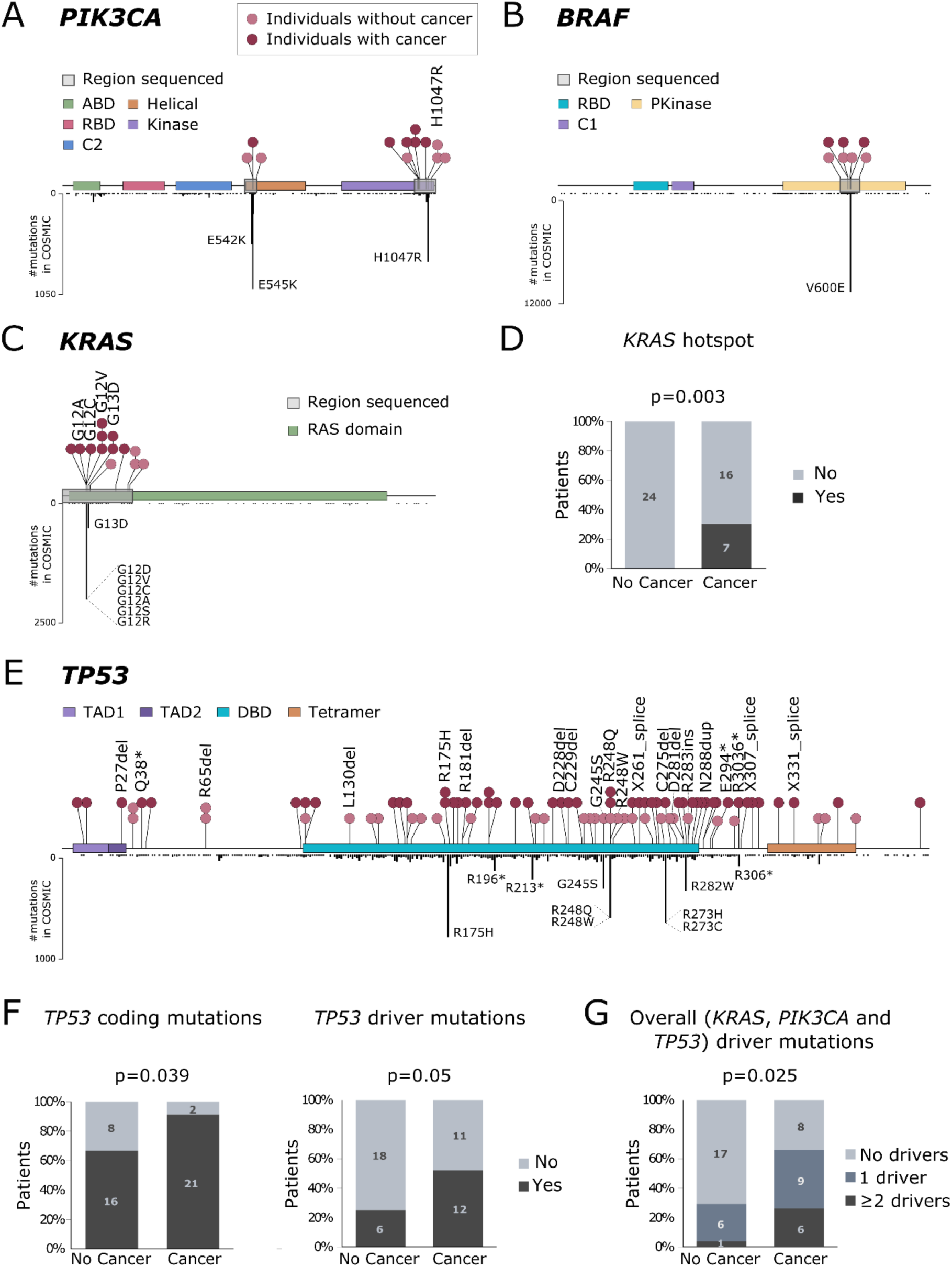
Normal colon carries mutations in common CRC genes, but these mutations are more abundant and pathogenic in patients with CRC. **A-C**. Distribution of mutations in *PIK3CA, BRAF* and *KRAS* in normal colon (above gene diagram) and in CRC samples from COSMIC database (below gene diagram). Normal colon mutations are color-coded by individuals with or without CRC and mutations corresponding to cancer hotspots are indicated. **D**. Percentage of patients with and without CRC that carry *KRAS* hotspot mutations in normal colon. **E**. Distribution of mutations across *TP53* in normal colon (above gene diagram) and in CRC samples from COSMIC database (below gene diagram). **F**. Percentage of patients with and without CRC that carry *TP53* coding mutations and driver mutations in their normal colon. **G**. Percentage of patients with and without CRC that carry one or more different cancer driver mutations in *PIK3CA, KRAS*, or *TP53* their normal colon. P-values correspond to Pearson Chi-Square. *ABD: adapter-binding domain; RBD: Ras-binding domain; Pkinase: protein tyrosine kinase domain; TAD: transactivation domain; DBD: DNA-binding domain; Tetramer: tetramerization domain*.

Given the tumor suppressor role of *TP53*, we deep sequenced all its coding exons. We identified a total of 85 coding mutations, which mostly clustered in the DNA binding domain of the protein and coinciding with areas of high density of CRC mutations in COSMIC (Fig. 3E and Supplementary Table S6). This clustering suggests that *TP53* mutations in normal colon are not random but follow patterns of selection similar to those operative in CRC. In addition, 17.7% of *TP53* mutations were high impact mutations (indels, nonsense, or splice), which severely affect protein function, and 9.4% of the substitutions identified corresponded to the top ten most common *TP53* substitutions in CRC, which represent >50% of all *TP53* mutations reported in COSMIC (Methods). In total, more than a quarter of *TP53* mutations identified in normal colon (27.1%) were either high impact mutations or hotspots. These mutations are likely to confer a selective advantage to the cells that carry them and were considered *TP53* driver mutations for the purpose of mutation classification in the study. Patients with CRC were more likely to carry *TP53* coding mutations in normal colon than patients without cancer (91.3% vs 66.7%, Pearson Chi-Square p=0.039, Fig. 3F) and also more likely to carry *TP53* driver mutations (52.2% vs 25%, Pearson Chi-Square p=0.05, Fig. 3F).

To investigate whether normal colon samples carried concurrently more than one cancer driver mutation, we plotted all the mutations identified for each gene in each patient (Fig. 4). In patients without CRC, we only observed one sample with multiple cancer driver mutations (2 in *TP53* and 1 in *PIK3CA*). However, in patients with CRC we identified 6 samples with multiple cancer driver mutations, including 4 samples with driver mutations in *KRAS* and *TP53*, and 2 samples with multiple driver mutations in *TP53*. Overall, patients with CRC were more likely to carry one or more cancer driver mutations in normal colon than patients without CRC (Pearson Chi-Square p=0.025, Fig. 3G). Thus, mutations in cancer driver genes are common in the normal colon of patients with CRC, not only as isolated events, but also in concurrence with other cancer driver mutations.

**Figure 4.**
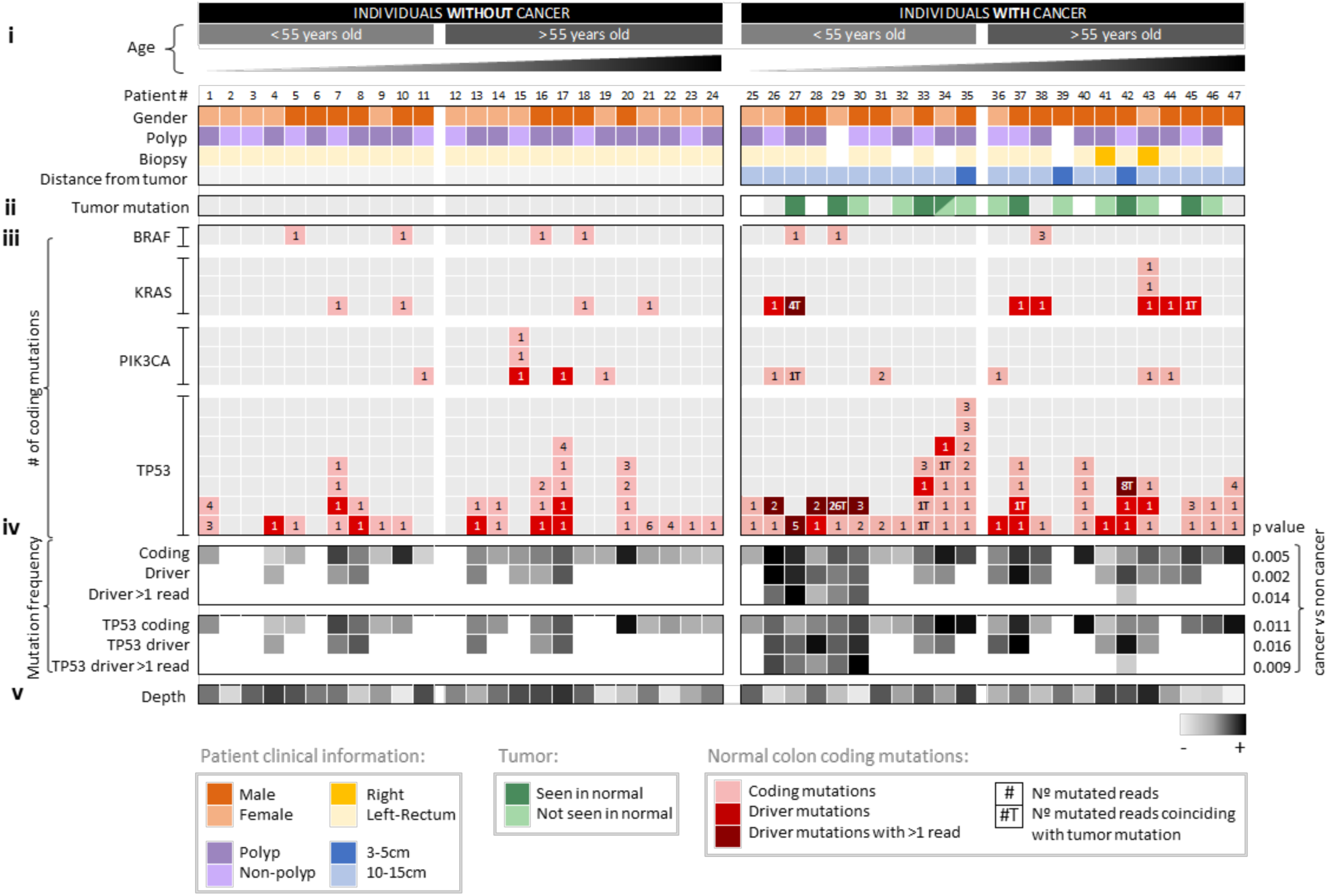
Mutations in normal colon of patients with CRC are often different from mutations in synchronous tumors and, in early onset CRC patients, driver mutations are frequently observed forming large clones. Each column corresponds to a patient. Patients are grouped by cancer status and sorted by ascending age. Panels of data indicate (**i**) clinical information, (**ii**) presence of tumor mutation in normal tissue, (**iii**) normal colon mutation counts for each gene, (**iv**) normal colon mutation frequency, and (**v**) depth. In (**i**) and (**ii**), white squares indicate that the information is not available and grey squares indicate negative. Tumor mutation was negative for four cases that did not show any mutation in the 4 tested genes. Normal colon mutations for each gene (**iii**) are indicated with squares that contain the number of mutated reads color coded for mutations that are coding, drivers, and drivers with more than one (>1) mutated duplex read. ‘T’ next to the number indicates that the mutation was observed in the synchronous tumor. Driver mutations were conservatively defined as oncogenic hotspots and *TP53* hotspots, nonsense, splice, and indel mutations. Greyscale heatmaps (**iv**) show mutation frequency values based on mutations that are coding, driver, and driver with >1 duplex read for all genes and *TP53* only. P-values correspond to t-test comparison of the mean frequency between individuals with and without CRC. Depth (**v**) indicates average duplex depth for all positions sequenced.

Of note, the MAF of the mutations identified, including driver mutations, was very low (<0.004 except for one mutation at 0.01) (Supplementary Fig. S4, Table S5, and Table S6), which makes them unidentifiable by standard sequencing methods (35). CRISPR-DS, however, enables accurate detection of very low frequency mutations, providing an ultra-high resolution view of the landscape of common CRC mutations in normal colon. The higher representation of cancer driver mutations in the normal colon of individuals with CRC indicates an excess of mutant clones in these patients whose detection by ultra-sensitive sequencing could be valuable for CRC risk prediction.

### Most mutations in normal colon of patients with CRC differ from mutations identified in the cancers of the same patients

We then investigated whether the mutations observed in the normal colon of individuals with CRC coincided with those detected in the synchronous tumor, indicating possible clonal origin. We sequenced the same 4 gene regions in tumor DNA from 19 patients with available tumor tissue and catalogued all non-synonymous mutations and indel mutations with MAF>0.1 (Supplementary Methods and Supplementary Table S7). In 4 tumors, no such mutations were identified, likely because they were driven by other non-sequenced genes. In 7 out of 15 tumors, at least one non-synonymous *TP53, KRAS* or *PIK3CA* mutation was identified in the tumor as well as the normal tissue (Fig. 4 and Supplementary Table S7). However, in all cases, the normal tissue also carried additional cancer gene mutations not detected in the tumor. In addition, in 8 cases, tumor *TP53, KRAS* and *PIK3CA* mutations could not be identified in the normal tissue, which nevertheless carried other mutations in these genes. Overall, out of 44 non-synonymous *TP53, KRAS, BRAF*, or *PIK3CA* mutations identified in the normal colon of 15 individuals with sequenced tumor data, only 9 (20.5%) coincided with a synchronous tumor mutation, indicating that most mutant clones observed in normal colon are not related to the clone that eventually progressed to CRC. These results suggest that multiple independent mutant clones might be abundant in normal colonic mucosa of individuals at risk of CRC, which increases the chance of eventually giving rise to a tumor.

While mutations in individuals without CRC were less abundant, in these patients we identified positive associations between the frequency of *TP53* coding mutations and being male (t-test p=0.056) and carrying polyps (t-test p=0.054) (Supplementary Fig. S5). Male gender and polyps are two well known risk factors for CRC (36,37). Smoking and high BMI are also CRC risk factors but were not associated with *TP53* mutation in patients without CRC in this study. These results suggest a potential link between *TP53* mutations in normal colon and some CRC risk factors.

### Clones with cancer driver mutations are larger in patients with early CRC

Next, we asked whether the mutations identified in the normal colon of individuals with CRC were not only more abundant but also corresponded to larger clones. As duplex depth indicates the number of haploid genomes sequenced, the number of duplex reads containing a given mutation is proportional to the relative size of the clone carrying the mutation. We observed that patients with CRC not only had more driver mutations, but these driver mutations were detected in multiple reads, indicating they were in larger clones (Fig. 4). Large clones were especially common in individuals that developed CRC younger than age 50. The largest clones observed corresponded to mutations also identified in the synchronous tumors, possibly pointing to very large clonal expansions or potentially dissemination of cancer cells more than 10cm from the tumor site. Overall, the frequency of coding, driver, and driver in more than one read mutations (for all genes and for *TP53* only) was statistically significantly higher in the normal colon of individuals with cancer compared to those without cancer (Fig. 4). However, the differences in mutation frequency between the two groups of patients (cancer and no cancer) were accentuated for younger (<55 yo) individuals (Supplementary Fig. S6). In addition, younger individuals with CRC carried a disproportional higher level of large mutant clones compared to older individuals with CRC (Supplementary Fig. S6), suggesting differences in the factors mediating clonal expansion and possibility progression to CRC in young and old individuals.

### *TP53* mutations in normal colon are more commonly pathogenic in individuals with CRC and more closely resemble mutations reported in CRC

*TP53* mutations were further assessed using Seshat, a web service tool that provides functional data specific for *TP53* variants, including frequency in the UMD database and predicted pathogenicity (38). We classified the mutations according to their location in the protein DNA-binding domain, their frequency in cancer, and their pathogenicity (See Supplementary Methods). Patients with CRC had a significantly higher mutation frequency of *TP53* mutations that are located in the DNA binding domain, are common in cancer, and are predicted to be pathogenic, compared to individuals without CRC (Fig. 5A).

**Figure 5.**
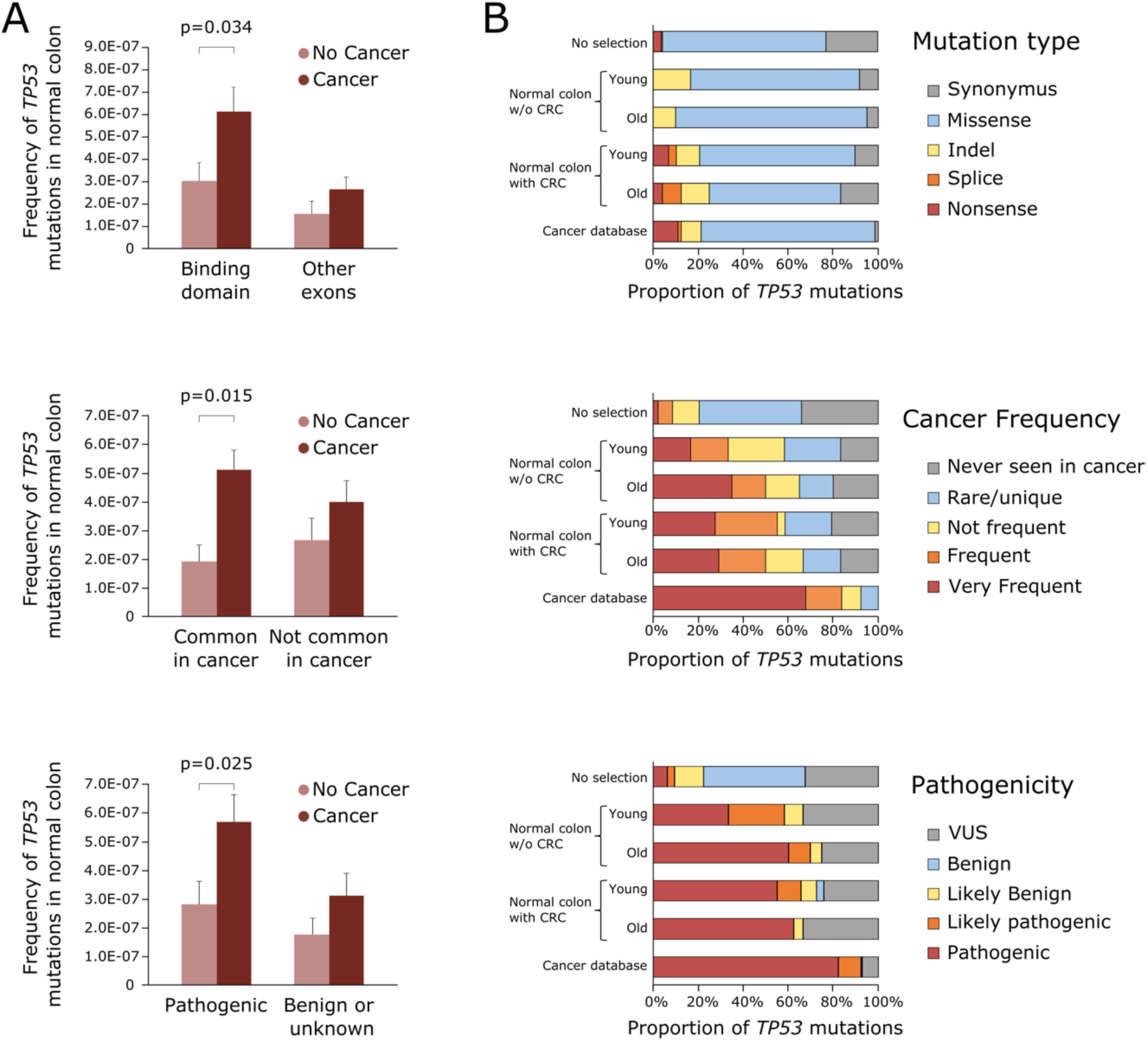
*TP53* mutations identified in normal colon are more pathogenic in individuals with CRC than individuals cancer-free and more closely resemble *TP53* mutations identified in CRC. **A**. *TP53* mutation frequency of individuals with and without CRC was compared based on mutations localized in the binding domain, mutations common in CRC, and mutations predicted to be pathogenic. Data was extracted from Seshat (ref. 38). Only significant p-values of t-tests are displayed. Error bars represent standard error of the mean. **B**. Distribution of *TP53* mutations by mutation type, cancer frequency, and pathogenicity in normal colon of young (<55 years old) and old (≥55 years old) individuals without and with CRC compared to all possible *TP53* mutations in the coding region (no selection, n= 3,546) and *TP53* mutations reported in CRC in the UMD cancer database (n=17,681). Number of *TP53* mutations in each group: young without CRC n=12; old without CRC n=20; young with CRC n=29; old with CRC n=24.

We then investigated the type, frequency, and pathogenicity of *TP53* mutations observed in normal colon compared with mutations reported in colon carcinomas in the UMD database (2021, n=17,681) and with all the possible *TP53* coding mutations in the theoretical absence of selection (n=3,546) (Fig. 5B). Mutations identified in normal colon samples were predominantly missense, similar to mutations reported in CRC or in *TP53* in the absence of selection. However, only the normal colon of patients with CRC carried nonsense and splicing mutations, which are considered highly damaging, in similar proportions to what is observed in the cancer database. The distribution of mutations reported in normal colon that are frequently found in cancers and predicted to be pathogenic clearly differed from the expected pattern of mutations in *TP53* under no selection and strongly resembled the pattern observed in the cancer database, especially in older individuals and those with CRC (Fig. 5B). These results suggest that there is a common process of positive selection of *TP53* mutant clones that is operative in normal colon as well as in CRC. However, this process appears to be enhanced with aging and in those patients that develop CRC.

### Integrative mutational analysis and proof-of-principle studies for the development of a CRC predictor

Our results provide support for the concept of creating a CRC risk predictor based on the normal colonic mucosa mutational profile. An essential step towards that goal is to construct a predictive model summarizing and harmonizing the mutational analysis results. Due to multicollinearity and to avoid overfitting, we used regularized logistic regression with Lasso penalty estimated to determine the 5 variables that were the best predictors (Supplementary Table S8, Methods). All quantitative variables with prior demonstrated significance in univariate analyses were included in the model as well as their interaction with age in order to determine potential differential effects between young and old individuals. The variables with the largest effects were the frequency of driver mutations (OR=2.16) and the presence of hotspots in *KRAS* (OR=1.86). Additional information was gained when considering the frequency of *TP53* coding mutations, *TP53* mutations common in cancer, and the interaction between drivers mutations with > 1 supporting read and age (ORs of 1.26, 1.066, and 1.26, respectively). This later interaction indicates that the risk of CRC increases with increased frequency of larger clones (represented by mutations identified in more than 1 read) but only in younger individuals. The predicted accuracy of the model was good, with AUC = 0.69 95% CI: 0.53-0.85 after 5-fold cross-validation. While this preliminary analysis included a small number of cases and requires validation in larger studies, it demonstrates the potential of this approach for the development of a CRC predictor based on the mutational analysis of biopsies collected from histologically normal mucosa.

## DISCUSSION

In this study we used deep sequencing with CRISPR-DS to perform high resolution single-molecule characterization of common colorectal cancer mutations in normal colon of individuals with and without CRC. We found that patients with CRC carried abundant oncogenic *KRAS* and *TP53* driver mutations in normal colonic epithelium distant from the primary tumor and that these clones were larger in patients with early onset CRC. In addition, most of the mutations identified in normal colon were different from the mutations in cancers, suggesting the presence of multiple, independent mutant clones in association with CRC progression. These results expand our understanding of somatic evolution in the colon, offer insights about different mechanisms of carcinogenesis in early vs late onset CRC, and raise the possibility of using normal colon biopsies for CRC risk assessment.

Prior studies have demonstrated an age associated increase of somatic mutations in normal colon (14,15,39) in agreement with our findings for non-coding mutations. By focusing on cancer driver mutations and leveraging ultra-deep, single-molecule sequencing, our study expands these initial findings and demonstrates that, in addition to the age-related increase of somatic mutations, the normal colon of individuals with CRC frequently carries clones with oncogenic *KRAS* and pathogenic *TP53* mutations. In some cases, we identified the same mutation in the normal biopsy and the tumor, which could be explained by a very large epithelial clonal expansion (at least 10cm long) from which the cancer evolved. Large clonal expansions have been previously described in the colon of patients with ulcerative colitis (40,41), who are prone to CRC, and exemplify the concept of carcinogenic fields (also known as field effect or field cancerization) by which the normal cell population is replaced by a cancer-primed cell population that is morphologically normal but carries some of the phenotypes required for malignancy (42). In most cases in our study, the driver mutation identified in normal colon did not coincide with the driver mutation in the tumor, suggesting the presence of fields composed by multiple precancerous clonal expansions as opposed to a single large clonal patch. Supporting this notion, six normal colon biopsies in patients with CRC carried two or more different driver mutations and, in three cases, one of the driver mutations was seen in the tumor, but not the others. While larger studies using multiple normal biopsies per individual are warranted to characterize the extension and composition of precancerous fields in CRC, the data obtained here reveals their high prevalence and demonstrates the feasibility of using single-molecule ultra-deep sequencing for their identification.

Lee-Six *et al* recently demonstrated that by the sixth decade of life around 1% of normal colon crypts carry a clonal driver mutation (15). Interestingly, most mutations identified were in genes rarely mutated in CRC, such as ERBB2 and ERBB3, and their frequency in normal colon was similar in patients with and without CRC. The authors suggested that these mutations likely contribute to crypt colonization by mutant stem cells whereas mutations in *TP53* and *KRAS* might enable subsequent preneoplastic transformation. Our results support this hypothesis by demonstrating that oncogenic *KRAS* mutations and pathogenic *TP53* mutations are infrequent in normal colon of patients without CRC but abundant in individuals that progressed to CRC. The finding of *KRAS* oncogenic mutations in normal colon of about one third of patients with CRC suggests a link between these mutant clones and the progression to CRC. *KRAS* mutations have been postulated to enable the lateral expansion of mutant crypts (43,44) providing a plausible mechanism for the generation of large patches of mutant cells from which tumors develop. In addition, phylogenetic reconstruction of CRC evolution has revealed that *TP53* and *KRAS* mutations are very early events, which take place after *APC* mutations decades prior to the development of CRC (1). Our results expand these findings by revealing that *TP53* and *KRAS* mutations can be identified even in histologically normal epithelium of patients with CRC suggesting that multiple mutant clones accumulate in these individuals through life, one of which eventually evolves into cancer.

In this study, we purposely included individuals with early onset CRC to investigate the nature of clonal expansions in the normal colon when cancer develops at younger age. The incidence of early onset CRC has increased substantially in the last 2 decades for unclear reasons (29). We discovered that almost one half of younger individuals with CRC (5/11) harbor a large clone containing a *TP53* driver mutation suggesting that the development of CRC in a large proportion of young individuals might be related to large expansions of *TP53* mutant clones in normal colon. Interestingly, early-onset CRC has been reported to have a high prevalence of *TP53* mutations and whole genome doubling (45), which suggests, in concordance to our data, that there might be different carcinogenic processes in early onset vs late onset CRC, the former involving frequent *TP53* loss in early stages of carcinogenesis.

In patients without CRC, we observed the highest levels of mutations in males and patients with polyps, which are risk factors for CRC. While larger numbers are required to confirm these associations, these findings agree with prior studies of somatic mutations in normal esophagus, in which smoking and alcohol consumption, which are well-known risk factors for esophageal adenocarcinoma, were associated with higher mutation burden (9). Thus, it is possible that the characterization of somatic mutation in normal colon might be useful to investigate the mechanisms of action of CRC epidemiological risk factors in addition to serve as a potential biomarker for cancer risk prediction.

A limitation of our study is that the gene panel was small and therefore, we cannot extract information about mutational signatures, which are helpful to elucidate mutation etiology. The goal of this study, however, was to determine whether ultra-sensitive sequencing of normal colon could have clinical value to assess CRC risk and, towards that goal, smaller panels are preferred to facilitate clinical applicability. In fact, we have demonstrated excellent predictive value by considering only the information provided by *KRAS* and *TP53* indicating that a future biomarker might be successful solely based on those genes. A smaller panel enables to dedicate sequencing resources to achieve higher depth and/or screen more biopsies, providing more accurate estimates of clone size and abundance. This approach is well aligned with recently proposed efforts to develop panels of hotspot cancer driver mutations as biomarkers of cancer risk using error corrected NGS (46). A second limitation of the study is that normal colon prior to the development of CRC was not tested. Moving forward, that assessment will be critical to demonstrate predictive value. The easy accessibility of the colon and established colonoscopic surveillance make it possible to obtain biopsies prior to CRC progression offering an excellent scenario for cancer risk prediction as well as a unique opportunity to study clonal evolution as a function of aging and exposures.

In summary, we have demonstrated that normal colon from patients with and without CRC carry mutations in common colorectal cancer genes, but these mutations are more abundant in patients with cancer. In addition, individuals with cancer carry more mutations that are canonical cancer drivers, especially in *KRAS* and *TP53*, and these mutations tend to be found in larger clones. Our results support the notion that somatic evolution contributes to clonal expansions in the normal colon and that this process is enhanced in individuals with cancer and, most significantly, in those with early onset CRC. These findings open the possibility for the development of a CRC predictor based on ultra-deep analysis of mutations in normal colonic biopsies.

## METHODS

### Subjects and samples

This study included normal colon mucosa samples (n=47) collected at the University of Washington Medical Center and affiliated practice sites (Seattle, WA, USA) from 24 patients without colorectal adenocarcinoma (CRC) undergoing colonoscopic screening or surveillance and from 23 patients with a newly diagnosed primary invasive colorectal adenocarcinoma undergoing surgical resection. Clinico-pathological characteristics of patients are listed in Supplementary Table S1. None of the patients had hereditary cancer syndrome. The groups of patients were matched by age and history of polyp(s) and were enriched with young individuals to explore differences in somatic mutations in early vs late onset CRC. Only one patient had neoadjuvant therapy. All normal samples from individuals with CRC were located 10 to 15 cm from the tumor except for two samples collected between 3 to 5 cm from the tumor. Immediately after collection, samples were frozen in liquid nitrogen and stored at -80ºC until DNA extraction (Supplementary Methods). In addition, Formalin-Fixed Paraffin-Embedded (FFPE) tumor blocks from patients with CRC were histologically examined with hematoxylin and eosin staining followed by microdissection and DNA extraction in 19 cases with sufficient tumor content. DNA extraction and library preparation from tumor DNA (Supplementary Methods) was performed after all normal tissue was analyzed to avoid any chances of cross-contamination. In all but one case, microsatellite instability (MSI) was determined by mismatch repair defect based on routine clinical immunohistochemistry of proteins MLH1, MSH2, MSH6, and PMS2 in tumor FFPE tissue sections. Two cases were MSI positive (Supplementary Table S1). Patients consented for sample collection and the study was conducted following protocols approved by Institutional Review Board committees at the University of Washington and the Fred Hutchinson Cancer Research Center. DNA from colorectal cancer cell lines HCT116, HT29 and SW480 was used for method validation (Supplementary Methods).

### CRISPR guide design

CRISPR-DS employs CRISPR-Cas9 digestion of target regions followed by size selection of excised fragments as a method for efficient target enrichment prior to library preparation (28) (Fig. 1B). We used Benchling [Biology Software, 2020] (San Francisco, CA) to design guide RNAs (gRNAs) to excise the coding regions of the *TP53* gene and the hotspot mutation codons of *BRAF, KRAS* and *PIK3CA* genes into fragments of ∼250-280bp. Then we used the CRISPOR web tool (47) to select the best candidates, which included 24 gRNAs (Supplementary Table S2) that excised the target region into 13 fragments with a total panel size of 3461bp. The panel comprised 1953 coding bp and 1508 non-coding bp from intronic regions flanking the excised exons.

### CRISPR-DS

Genomic DNA from normal colon tissues and CRC cell lines was processed for CRISPR-DS as previously described with minor modifications (28) (Supplementary Methods). Hybridization capture was performed with 120bp biotinylated xGen Lockdown probes (Integrated DNA Technology, Coralville, IA, USA) designed to target the selected regions of *TP53, BRAF, KRAS* and *PIK3CA* (Supplementary Table S3). Libraries were sequenced using 150 PE reads on a MiSeq Illumina platform on site or HiSeq at Genewiz (South Plainfield, NJ), allocating ∼2 million reads per sample. Sequencing reads were analyzed as previously described (28) using pipeline v1.1.4 from https://github.com/Kennedy-Lab-UW/Duplex-Seq-Pipeline (Supplementary Methods). Mutant Allele Frequency (MAF) was calculated for each mutation as the number of mutated duplex reads divided by the total duplex depth at the given position.

### Calculation of mutation frequency

For each sample, the overall duplex depth was calculated as the total number of duplex nucleotides sequenced divided by the size of the panel. On average, for each sample we sequenced 8.6M duplex nucleotides corresponding to a duplex depth of 2,484x (minimum 1,268x; maximum 4,306x) (Supplementary Table S4). To correct for the variability in sequencing depth across samples (Fig. S2), sample comparisons were made based on mutation frequencies, which were calculated as the number of mutations in a given region (e.g., coding, non-coding, *TP53* coding) divided by the total number of duplex nucleotides sequenced in that region. Coding included nucleotides in coding exons plus 2bp boundary nucleotides to capture splice site mutations, and non-coding included all the remaining nucleotides in the target regions. Similarly, mutation frequencies were calculated for specific types of mutations (e.g., drivers) by dividing the number of mutations in the category of interest by the total number of duplex nucleotides sequenced in the target region. Mutation counts and corresponding mutation frequencies for each sample are indicated in Supplementary Table S4.

### Mutational analysis

Coding mutations were extracted from MAF files (Supplementary Methods) and were further annotated by mutation type (missense, nonsense, splice, indel and synonymous), mutation spectrum (C>A, C>G, C>T, T>A, T>C and T>G), localization in CpG dinucleotides, and driver mutations. Mutations in *BRAF, KRAS*, and *PIK3CA* were considered driver mutations if they corresponded to common oncogenic hotspot mutations in these genes according to large intestine carcinoma data from the COSMIC database (https://cancer.sanger.ac.uk/cosmic) (34). The following oncogenic driver mutations were considered: *BRAF* V600E, which accounts for >90% of *BRAF* mutations in CRC; *KRAS* hotspot mutations in codons 12 and 13, which account for >90% of *KRAS* mutations in CRC; and *PIK3CA* hotspot mutations E545K, H1047R and E542K, which account for >50% of *PIK3CA* mutations in CRC. Driver mutations in *TP53* included the 10 most common substitutions according to the COSMIC database, which represent >50% of all mutations reported in large intestine carcinomas (p.R175H, p.R273H, p.R248Q, p.R282W, p.R273C, p.R248W, p.G245S, p.R213*, p.R196* and p.R306*), and all splice, indels and nonsense mutations. The list of annotated coding mutations for oncogenes and *TP53* are presented in Supplementary Tables S5 and S6, respectively. Large intestine carcinoma variants from COSMIC were also used to determine the mutation spectrum (6 possible nucleotide substitutions) of CRC (n=70,525) (Fig. 2E) as well as the distribution of CRC mutations within the protein domains of the genes of interest (Fig. 3A-C, E).

### Statistical analysis

Correlations were tested with Spearman’s rank test. Comparison of mutation frequency means across groups of individuals was performed by t-test. Associations between categorical variables were tested with Pearson Chi-Square. All tests were two-sided at an alpha level (type 1 error rate) of 0.05. The predictive model was estimated with the glmnet R package (48), with parameters for Lasso logistic regression. The penalization parameter was selected to restrict the model to 5 covariates. Predictive accuracy was calculated with the area under the ROC curve and its 95% confidence intervals as implemented in the pROC R package (49). Statistical analyses were performed with SPSS version 25 and R version 3.6.3.

## Data Availability

All data produced in the present study are available upon reasonable request to the authors

## Software availability

Software is available at https://github.com/Kennedy-Lab-UW/Duplex-Seq-Pipeline.

## Data access

Sequencing data from this study have been submitted to the NCBI BioProject database (https://www.ncbi.nlm.nih.gov/bioproject) under accession number PRJNA767868.

## Acknowledgments

This work was supported in part by i-PFIs (IFI16/00012) and M-AES (MV19/00042) research fellowships from Instituto de Salud Carlos III to JM; Agencia Estatal de Investigación (RTI2018-094009-B-100) to MAP; startup funds from the Department of Laboratory Medicine and Pathology to RAR; R50CA233042 to MY; P01CA077852, P30CA042014, P30CA15704, R01189184, R01CA194663, R01CA207371, R01CA220004, U01CA206110, U54CA143862, Listwin Family Foundation, Cottrell Family Fund, R.A.C.E.Charities, Seattle Translational Tumor Research, Rodger C. Haggitt Endowed Chair, and W.H. Geiger Family Foundation to WMG; and Agency for Management of University and Research Grants (AGAUR) of the Catalan Government grant 2017SGR723 to VM. We thank Dr. Scott Kennedy for manuscript revision and members of the Risques and Kennedy Lab at the University of Washington and Grady lab at the Fred Hutchinson Cancer Research Center for technical assistance. We are grateful to the ColoCare and GiCare Studies for providing patient samples and we deeply thank all the patients who donated samples, without which this research would not have been possible.

## Author Contributions

JM, WG, MAP and RAR designed research, selected tissues to be studied, and acquired tissue samples; JM, JF, and KC performed research; JM, BK, TW, VM, and RAR analyzed the data; SG performed pathological analysis; TS provided analytical resources; MY supervised sample selection; JM, WG, MAP and RAR interpreted data; JM and RAR wrote the article.

## SUPPLEMENTARY METHODS

### DNA extraction

Genomic DNA was extracted from frozen normal colon tissue samples using the DNeasy Blood & Tissue Kit (Qiagen, Hilden, Germany) and from the FFPE tumor samples using the QIAamp DNA FFPE Tissue Kit (Qiagen). Forceps mucosal biopsies procured at endoscopy were approximately 6mm x 4mm x 3mm in size and the whole biopsy was used for DNA extraction. In normal colon samples from surgical resections, mucosal epithelium was selected to match the size of the endoscopic biopsies. DNA was quantified by Quant-iT PicoGreen dsDNA Assay Kit (Life Technologies). Cell line DNA was extracted using PureLink™ Genomic DNA Mini Kit (Invitrogen, Waltham, MA) and DNA was quantified by Qubit dsDNA BR Assay Kit (ThermoFisher Scientific, Waltham, MA). For cell lines and a subset of normal colon samples, DNA quality was assessed with Genomic TapeStation (Agilent Technologies, Santa Clara, CA) and demonstrated high quality in all samples (DNA integrity number (DIN) ≥7).

### CRISPR-DS library preparation

CRISPR-DS was performed according to our published protocol (1). Briefly, 100ng of DNA were digested with gRNA-Cas9 complex followed by double size selection with AMPure XP Beads (Beckman Coulter, Brea, CA, USA) to enrich for ∼260bp on-target fragments and remove uncut genomic DNA. Then, DNA fragments were end-repaired, A-tailed and ligated with duplex adapters containing 8 bp random double-stranded molecular tags (TwinStrand Biosciences, Seattle, WA, USA) using the NEBNext Ultra II DNA Library Prep Kit (NEB, Ipswich, MA, USA). Ligated DNA was amplified using the KAPA Real-Time Amplification kit with fluorescent standards (KAPA Biosystems, Woburn, MA, USA). Upon purification, regions of interest were captured by hybridization with 120bp biotinylated xGen Lockdown probes designed to capture previously excised regions of the genes of interest (Integrated DNA Technology, Coralville, IA, USA) (Table S3). A post-capture PCR using indexed primers was performed with KAPA HiFi HotStart PCR kit (KAPA Biosystems, Woburn, MA, USA). Libraries were visualized on the Agilent 4200 TapeStation to confirm the expected peak size. If peaks were not present, a second round of hybridization capture was performed. Libraries were quantified using the Qubit dsDNA HS Assay kit, diluted, and pooled for sequencing.

### CRISPR-DS data analysis

CRISPR-DS sequencing data was analyzed as previously described (1) using pipeline v1.1.4 available at https://github.com/Kennedy-Lab-UW/Duplex-Seq-Pipeline. First, raw reads were grouped using the double stranded molecular tag included in the duplex adapters and a Single-Strand Consensus Sequence read was built from reads sharing the same tag. Then Single-Strand Consensus Sequence reads with complementary tags were compared to produce a single, highly accurate duplex read (Fig. 1B). Duplex reads were aligned to the human genome reference hg38 (GRCH38), end-trimmed, locally realigned, and overlap-trimmed. Variants were called using a samtools mpileup-based variant caller and output VCF files were converted to MAF files using the Vcf2Maf script (https://github.com/mskcc/vcf2maf) with VEP version 99. All variants identified in SNP positions were discarded for subsequent analysis. Three samples with potential cross-contamination based on SNP frequency were removed from the study. For each mutation, Mutant Allele Frequency (MAF) was calculated as the number of mutated duplex reads divided by the total duplex depth at the given position.

### CRISPR-DS validation using CRC cell lines

CRISPR-DS reproducibility, sensitivity, and efficiency were evaluated using DNA extracted from three common human colorectal cancer cell lines obtained from ATCC (HCT116, HT29 and SW480). One hundred (100) ng of DNA from each cell line was processed for CRISPR-DS in two independent replicate libraries to test reproducibility. To test sensitivity, HT29 DNA was spiked in HCT116 DNA at three different ratios (1:10, 1:20 and 1:100). Library preparation and data analysis were performed using the same methods employed for tissue samples. As each duplex read corresponds to an original DNA molecule, duplex depth indicates the number of haploid genomes analyzed in each position. Thus, the efficiency (also called recovery rate) was calculated as the average duplex depth divided by the number of input genomes corresponding to 100ng of DNA. Driver mutations in each cell line (HCT116: *KRAS* G13A and *PIK3CA* H1047A; SW480: *KRAS* G12V, *TP53* R273H and P309S; HT29: *BRAF* V600E and *TP53* R273H; based on https://web.expasy.org/cellosaurus/) were identified in all replicates and with high sensitivity in the spike-in experiment, validating the assay (Supplementary Fig. S1).

### Tumor Sequencing

50-100ng of tumor FFPE DNA were sonicated, end repaired, A-tailed and ligated to duplex adapters using commercial kits (TwinStrand Biosciences, Seattle, WA). Hybridization capture was performed with the same probe pool used for CRISPR-DS of normal colon but using two rounds of hybridization capture as previously recommended (2) (Supplementary Table S3). Enriched libraries were amplified, quantified using the Qubit dsDNA HS Assay kit, diluted, and pooled for sequencing on a MiSeq Illumina platform on site allocating ∼0.8 million reads per sample. Raw reads were processed with the DS pipeline v2.1.2 available at https://github.com/Kennedy-Lab-UW/Duplex-Seq-Pipeline. Data analysis was performed on Single-Strand Consensus Sequence (SSCS) reads instead of duplex reads to provide higher depth (mean 284x) and because the goal was to identify tumor driver mutations, which are expected to be clonal and harbor large MAF. For each tumor, we catalogued *BRAF, KRAS, PIK3CA*, and *TP53* non-synonymous or indel mutations with MAF>0.1 and determined whether these mutations coincided with mutations identified in normal colon from the same individual.

### *TP53* mutation characterization with Seshat

*TP53* mutations were characterized using the Seshat web service tool (https://p53.fr/TP53-database/seshat) (3). A MAF file containing all the *TP53* mutations observed (n=118) was submitted to Seshat to accurately annotate, validate, and analyze *TP53* variants using data derived from the UMD *TP53* database (4). From the Seshat output, coding and splice mutations were extracted (n=85) (Table S6) along with the information about their frequency in the UMD cancer database and predicted pathogenicity. Frequency in the cancer database was categorized as “Common in cancer” (including very frequent and frequent mutations) and “Not common in cancer” (including not frequent, rare, unique, and not seen before mutations). Predicted pathogenicity was categorized as “Pathogenic” (including pathogenic and likely pathogenic mutations) and “Benign or unknown” (including likely benign, benign and variants of unknown significance). In addition, the distribution of TP53 mutations in normal colon based on mutation type, cancer frequency, and pathogenicity was compared to the expected distribution for random *TP53* mutations as well as CRC *TP53* mutations based on the UMD database (Supplementary Methods).

### *TP53* mutations without selection

To compare *TP53* mutations in normal colon with the theoretical make up of mutations if they were to occur completely at random and without selection, we generated a list of all possible mutations in the *TP53* coding region (n=3,546) and submitted it to Seshat to determine their frequency in cancer and predicted pathogenicity.

### UMD *TP53* cancer database mutational analysis

To compare *TP53* mutations observed in normal colon with the mutations present in CRC, we used the most recent UMD *TP53* cancer database (2021) kindly provided by Dr. Thierry Soussi (Sorbonne Université, Paris, France). We selected all mutations corresponding to colorectal carcinoma samples (n=17,681) and determined the distribution of mutations according to mutation type, frequency in CRC, and predicted pathogenicity. The distribution of *TP53* CRC mutations across these variables was compared with the distribution of normal colon mutations for the same variables, divided by patients younger and older than 55 years old and patients with and without CRC.

### Bi-Sulfite Conversion and methylation assessment

DNA (500 ng) from each sample was bisulfite converted using the EZ DNA Methylation Kit (Zymo Research, Irvine, USA). The DNA samples were submitted to the Genomics Core at the Fred Hutchinson Cancer Research Center where they were processed and run on MethylationEPIC arrays following the manufacturer’s instructions (Illumina, Inc., San Diego, CA). As previously described, the raw intensity files (IDAT) were preprocessed, normalized, and the array results assessed using four epigenetic age clocks (Hannum, Horvath, PhenoAge and EpiTOC (5)).

## SUPPLEMENTARY FIGURES

**Figure S1.**
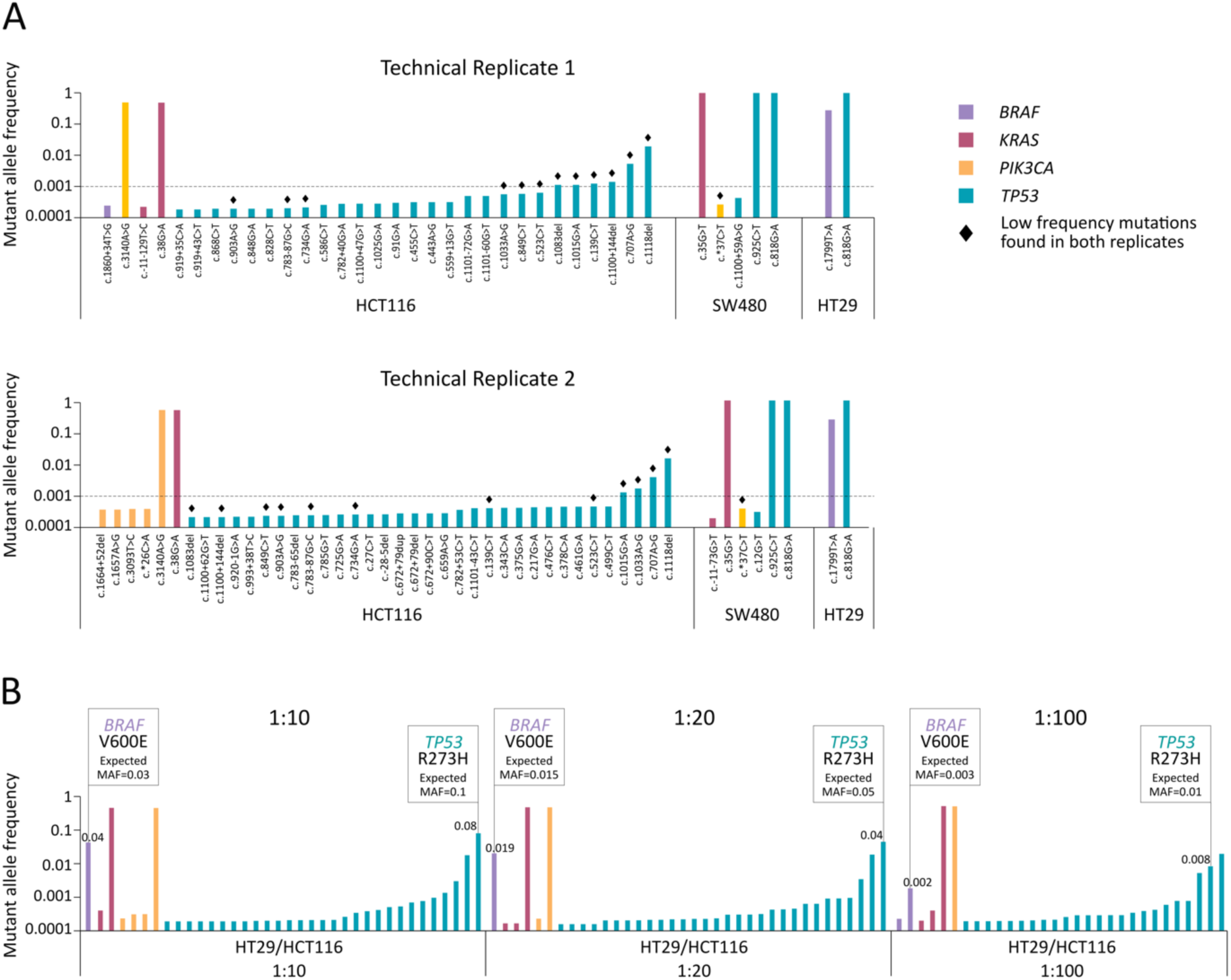
Ultra-deep sequencing of colorectal cancer cell lines with CRISPR-DS. **A**. Distribution of mutations in two technical replicates of HCT116, SW480 and HT29 cell lines. Each bar represents a mutation, color coded by gene. The height of the bars indicates Mutant Allele Frequency (MAF), calculated as the number of mutant alleles divided by the sequencing depth at a given position. Mutations are sorted by ascending MAF within each gene for each sample. Mutations found in both replicates are highlighted with a black diamond. **B**. Spike in of HT29 in HCT116 at different concentrations: 1/10, 1/20 and 1/100. Expected HT29 mutations and their MAF for each dilution are indicated with boxes and the observed frequency is indicated above each of the corresponding bars.

**Figure S2.**
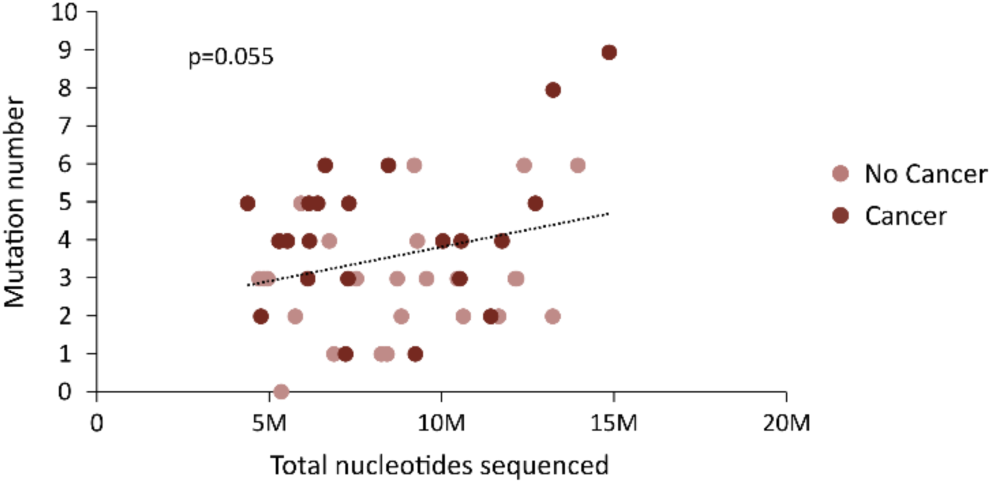
Number of mutations tends to increase with number of total nucleotides sequenced. Correlation between the number of mutations and the total nucleotides sequenced by patient. P-value corresponds to Pearson correlation.

**Figure S3.**
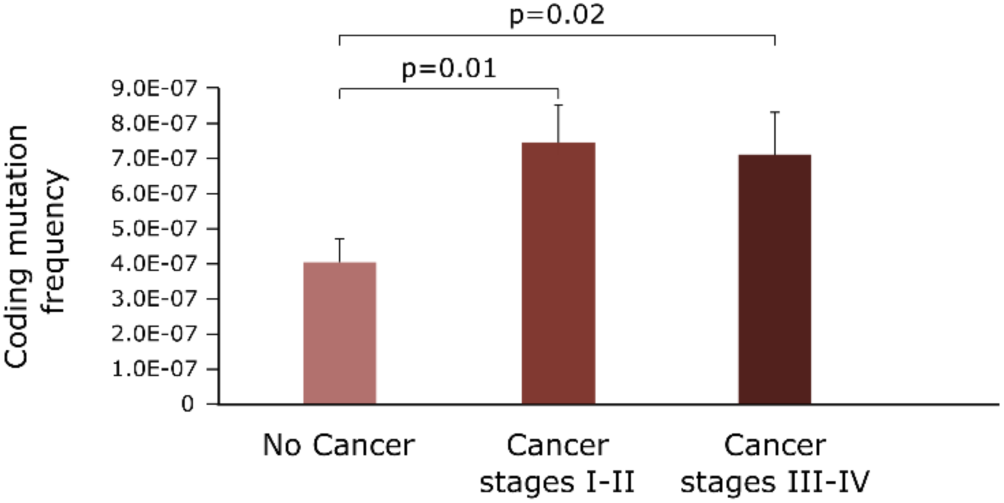
Normal colon from individuals with CRC carries higher coding mutation frequency than individuals without cancer regardless of the cancer being early or late stage. Coding mutation frequency is calculated as the number of mutations divided by the total number of duplex nucleotides sequenced in the coding target regions. P-value corresponds to t-test. Error bars represent standard error of the mean.

**Figure S4.**
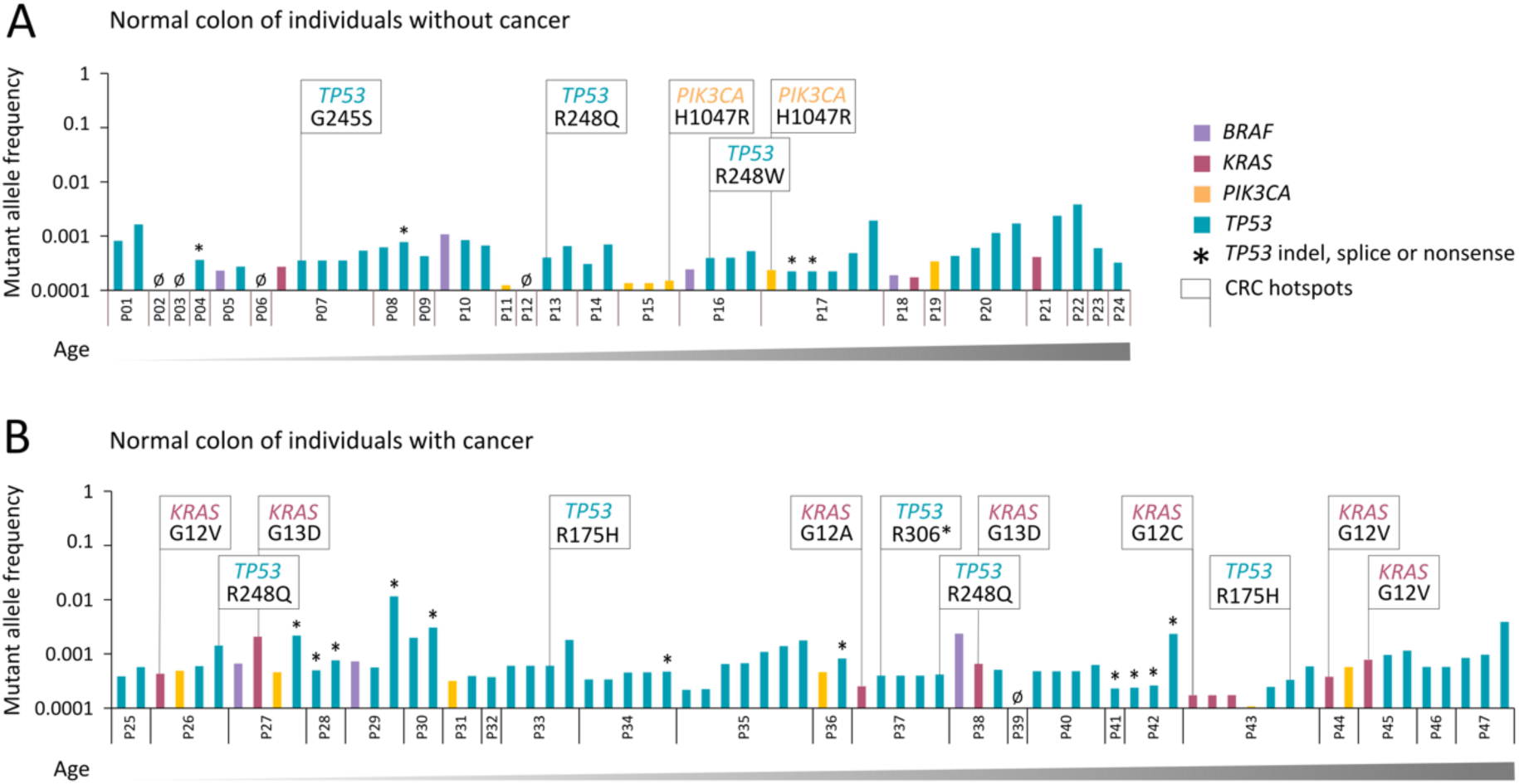
Normal colon from individuals with and without CRC carries mutations in common cancer genes. Distribution of mutations in normal colon from individuals without CRC **(A)** and individuals with CRC **(B)**. Patients are sorted by ascending age and patient IDs are indicated in the x-axis. Each bar represents a mutation, color coded by gene. The height of the bars indicates Mutant Allele Frequency (MAF), calculated as the number of mutant alleles divided by the sequencing depth at a given position. Mutations are sorted by ascending MAF within each gene for each patient. Hotspot mutations are indicated with boxes and *TP53* indels, nonsense, and splice mutations are indicated with asterisks.

**Figure S5.**
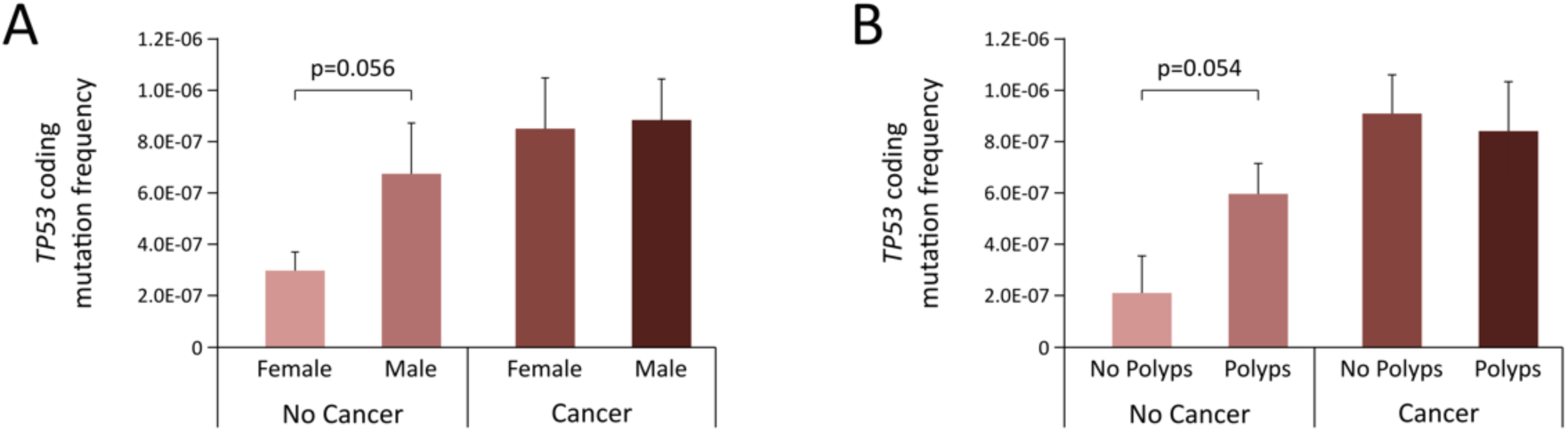
*TP53* coding mutations are more frequent in normal colon from individuals without CRC that are males or harbor polyps. Comparison of *TP53* mutation frequency in the normal colon of (**A**) females and males with and without CRC and (**B**) polyp and non-polyp formers with and without CRC. P-values correspond to t-test comparisons. Error bars represent standard error of the mean.

**Figure S6.**
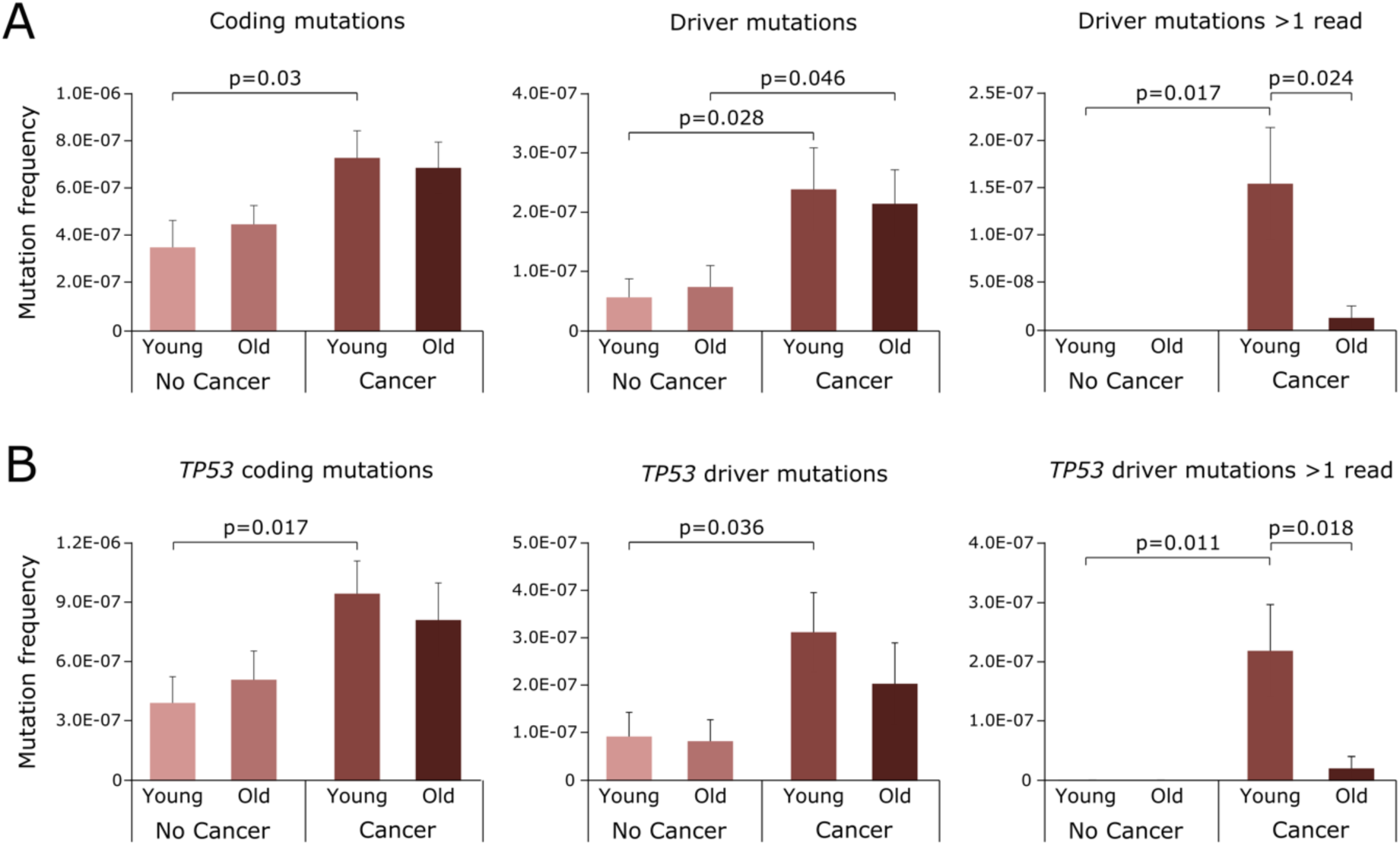
Driver mutations and larger clones are more abundant in the normal colon of young patients with CRC. Comparison of mutation frequency in the normal colon of younger (<55 year old) and older (≥55 year old) individuals with and without CRC. Mutation frequencies were calculated for overall (**A**) and *TP53*-only (**B**) mutations that are coding, drivers, and drivers with >1 mutated duplex read. P-values correspond to t-test comparisons. Error bars represent standard error of the mean.

